# Laboratory-based surveillance of non-tuberculous mycobacterial pulmonary disease in Japan

**DOI:** 10.1101/2024.04.02.24305177

**Authors:** Yuko Hamaguchi, Kozo Morimoto, Satoshi Mitarai

## Abstract

**Background:** Intractable pulmonary diseases with non-tuberculous mycobacteria (NTM-PDs) and antimicrobial resistance have become increasingly concerning worldwide. Nevertheless, a surveillance system for NTM has not been established in most countries, thus requiring repeated, intermittent, and time-consuming cross-sectional nationwide surveys.

**Methods:** To establish a nationwide surveillance system for NTM-associated diseases, we aimed to develop a prototype computer system primarily designed to continuously estimate NTM-PD incidence using a bacteriological case-defining algorithm through an automatic process of integrating bacteriological data collected from microbiology laboratories across Japan. To validate the accuracy of our study results, we compared the distribution of pulmonary TB cases between our laboratory data and the national surveillance data, which is representative of the Japanese population.

**Results:** Our estimates implied a 17.7% increase in NTM-PD incidence from 15.8 [14.9–16.8] in 2013 to 19.2 [18.2–20.4] in 2017, per 100,000 population. Moreover, 93.0% of the identified NTM-PD cases were dominated by the *Mycobacterium avium*-*intracellulare* complex, and the proportion of *M. abscessus* species exceeded that of *M. kansasii* for the first time in Japan. We also revealed significant age and sex differences in NTM-PDs. Notably, we found similar characteristics between our laboratory data and national surveillance data covering almost the entire Japanese population.

**Conclusion:** These findings, despite being laboratory-based, are extrapolatable to the general population in Japan and provide evidence that supports our system as a viable alternative to the nationwide NTM surveillance system.

## 1. Introduction

Pulmonary diseases caused by non-tuberculous mycobacteria (NTM-PDs) have become increasingly concerning worldwide. Studies in the 2000s have indicated that the prevalence of NTM-PDs was higher in Japan than in industrialised countries such as the United States, Denmark, France, Greece, Australia, and New Zealand [1-3] Since the first nationwide survey conducted in 13 tuberculosis (TB) sanatoriums in Japan in 1968, nationwide surveys have been employed to estimate the NTM-PD incidence using a unified formula between 1971 and 1997 (Supplementary Figure S1) [4]. Since then, the incidence of NTM-PDs has been estimated in nationwide cross-sectional surveys conducted intermittently in 2001, 2007, and 2014 [5]. These studies have shown that although the incidence of pulmonary TB has decreased, the incidence of NTM-PDs has been gradually increasing since 1971 and peaked at 14.7 in 2014, surpassing the pulmonary TB notification rate of 11.9 (Supplementary Figure S1). *Mycobacterium avium, M. intracellulare, M. kansasii*, followed by *M. abscessus*, are the most isolated species from patients with NTM-PD. Many of these cases, excluding those caused by *M. kansasii*, exhibit resistance to chemotherapy and are highly prone to recurrence [5].

The extent to which such intractable NTM-associated diseases may spread nationwide should be seriously considered for public health. Nevertheless, comprehensive nationwide notification and population-based surveillance systems for NTM are lacking in most countries, including Japan. Consequently, there is a reliance on conducting nationwide cross-sectional surveys. However, despite great costs and efforts, very limited information has been obtained [3, 6, 7]. Therefore, it is essential to develop a population-based surveillance system for NTM that integrates individual clinical and bacteriological data. Such a system is essential for answering numerous research questions related to NTM-PDs and for advancing our understanding and management of these diseases.

Private laboratories in Japan cover more than 80% of laboratory tests outsourced from medical facilities nationwide; thus, a massive amount of bacteriological information related to the diagnosis of TB or NTM-associated diseases has been amassed over time. Each examinee is linked to their individual data, geographic information, and examination status (e.g. examined specimen, culture testing, and genetic analysis with nucleic acid amplification testing (NAT), DNA-DNA hybridisation (DDH), and matrix-assisted laser desorption/ionisation time of flight mass spectrometry (MALDI-TOF MS). Our recent study [8] presented preliminary findings on the two-year period prevalence and proportional data by species using bacteriological information from private laboratory data, as mentioned above. Nevertheless, owing to the lack of an observational period, we could not estimate the annual NTM incidence rates. Therefore, this study aimed to 1) develop a preliminary database system that integrates bacteriological information from private laboratories across Japan; 2) design computer algorithms of bacteriological criteria defining respiratory infections with NTM (or TB); and 3) continuously estimate the annual NTM-PD incidence.

## 2. Material and methods

### 2.1. Data collection

#### 2.1.1. Mycobacterial data in major microbiology laboratories

Two of the predominant private laboratories in Tokyo, Japan, SRL, Inc. and LSI Medience Corporation, from which most medical facilities across Japan outsource bacterial examinations, offered laboratory mycobacterial examination data. The data consisted of cross-sectional and chronological data of individuals (names were anonymized by mixing symbolic letters) who were registered as examinees between 2012 and 2020. The data included the following information: date when specimens were obtained from the examinees, registered, and examined in the laboratories; age; sex; medical facilities and locations (prefectures) where the specimens were obtained; specimens (e.g. sputum, bronchoalveolar lavage fluid, transbronchial lung biopsy); and examination status (culture test, NAT, DDH, and MALDI-TOF MS).

#### 2.1.2. TB notification data

Annual reports of TB statistics (all forms except latent TB infection) were collected from the Tuberculosis Surveillance Center at the Research Institute of Tuberculosis, which was established to manage TB surveillance as part of the national surveillance system (National Epidemiological Surveillance of Infectious Diseases) [9].

#### 2.1.3. Population data

Annual population statistics, including estimates and population censuses (conducted once every five years), were obtained from the Statistics Bureau of Japan [10]. We reconstructed a new dataset based on year, age, sex, prefecture, and region to calculate and estimate the incidence of pulmonary TB and NTM-PD.

### 2.2. Case definition

#### 2.2.1. Case identification

To identify examinees, unique identification codes (unique-ID) were created. These consisted of age, sex, laboratory code (two private laboratory branches located near the medical facilities), medical facility code, and an anonymised code of each examinee.

#### 2.2.2. Exclusion criteria to correct for bias due to censoring data

As NTM-PD cases were more likely to be diagnosed within one to two years (mostly less than one year) after the first consultation, the incidence rate in the first year after registration could be overestimated. Therefore, we excluded all examinees in the first year after registration. We also excluded all examinees who were registered after the final two years of the entire registration period as censoring cases, as they could not complete two years of follow-up.

#### 2.2.3. Bacteriological diagnostic criterion for NTM-PD and pulmonary TB

To define NTM-PD cases, we referred to the bacteriological criteria, which are part of the NTM-PD diagnostic criterion in an official clinical practice guideline updated in 2020 by the American Thoracic Society (ATS), European Respiratory Society (ERS), European Society of Clinical Microbiology and Infectious Diseases (ESCMID), and Infectious Diseases Society of America (IDSA) [11]. The ATS/ERS/ESCMID/IDSA diagnostic criteria include the following: 1) positive culture results from sputum specimens expectorated on at least two separate dates or 2) at least one positive culture result from bronchial wash, lavage, transbronchial, or other lung biopsy showing mycobacterial histologic features. Thus, we defined the examinees as NTM-PD cases if they satisfied the above bacteriological criteria within a follow-up period of two years since they were first registered (Supplementary Figure S2). Pulmonary TB cases were also identified according to bacteriological criteria in which respiratory or gastric fluid specimens tested positive for NAT at least once (Supplementary Figure S2).

### 2.3. Analysis

#### 2.3.1. Basic statistics

Basic statistics, including age and sex, were calculated to summarise the characteristics of the sample population. The 95% confidence intervals (CI) were calculated assuming that the incidence of NTM-PD and pulmonary TB cases followed Poisson distribution. The NTM species isolated and identified from the culture-positive samples were also summarised.

#### 2.3.2. Estimates of the incidence rate of NTM-PD

To analyse the epidemic dynamics of NTM-PD, the annual incidence rates (per 100,000) and number of NTM-PD cases between 2013 and 2017 were estimated using the same formula (Supplementary Table S1), as per previous studies [8, 12].

#### 2.3.3. Statistical analysis

The Cochran–Armitage trend test was performed to evaluate the time- and age-dependent trends in NTM-PD incidence. The chi-square test was used to evaluate sex differences in the incidence of NTM-PD. Differences in the mean ages between females and males were statistically tested using the Wilcoxon rank-sum test (compared to the Welch two-sample t-test).

### 2.4. Ethical approval

Bacteriological data were anonymized to protect privacy before collection, except for analysis of minimal and essential information, such as age, sex, and prefectures where the medical facilities accessed by the examinees were located. This study was approved by the Ethics Committee of Fukujuji Hospital (approval #19046). The need to obtain the informed consent was waived due to the retrospective nature of the study.

## 3. Results

### 3.1 Case enrolment and bacteriological and TB surveillance data alignment

Our dataset included 363,163 samples from 449,583 cases, integrating 131,133 samples from LSI and 232,030 samples from SRL. Following the exclusion of ineligible cases (200,949), a total of 21,791 NTM-PD and 14,420 pulmonary TB cases satisfied the bacteriological criteria mentioned above (Figure 1). A comparison of age and sex distribution between pulmonary TB cases in our study population and those in the national TB notifications indicated similarity; the former had a mean age of 71 (95% CI; 70.8–72.1) with 36.4% females and the latter had a mean age of 67 (95% CI; 67.0– 67.5) with 36.7% females (Table 1 and Supplementary Figure S3). This indicated that the sample population used in this study was likely similar to the nationally representative TB surveillance data.

**Figure 1.**
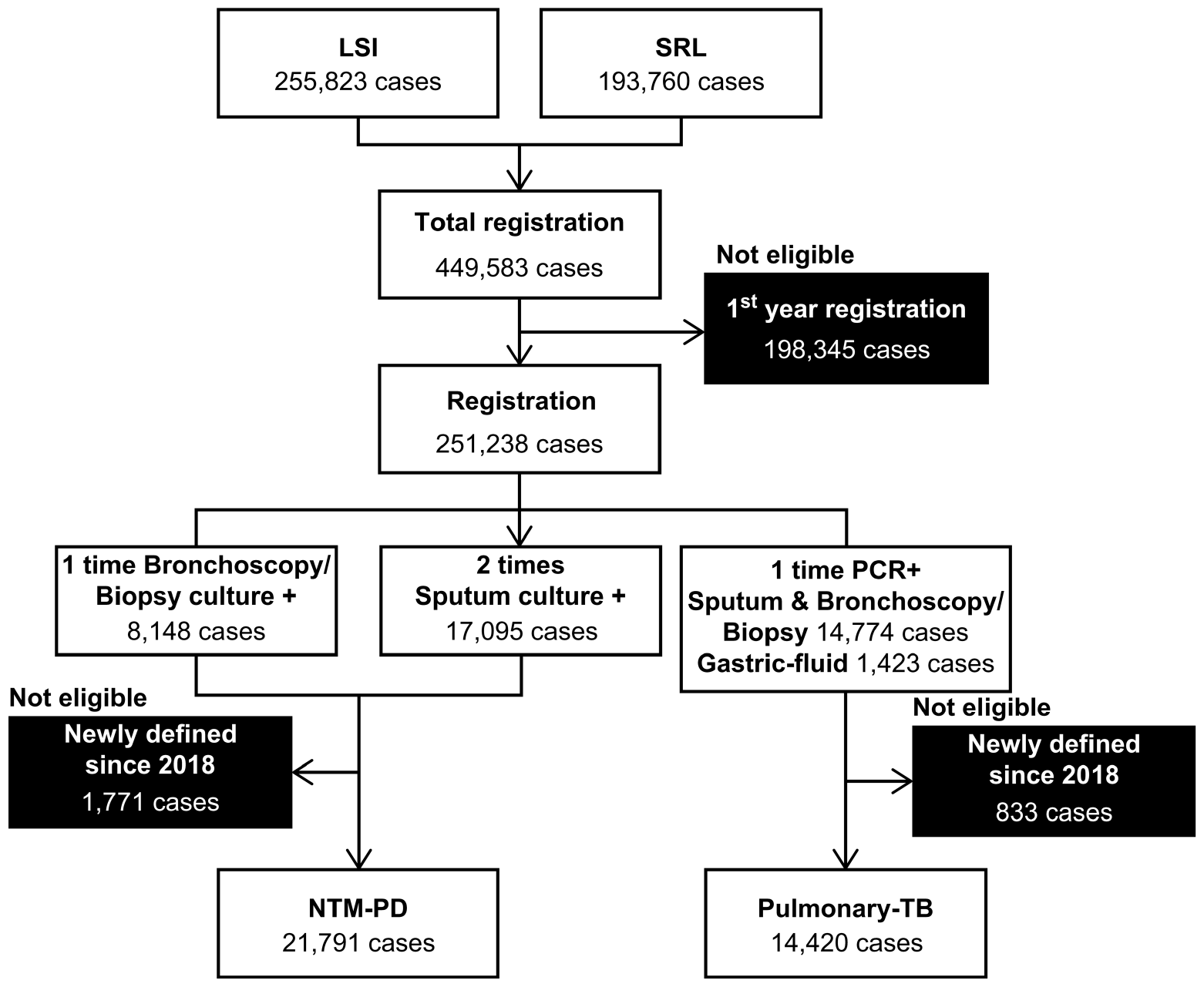
Flow chart of bacteriological criteria selection for case enrolment.

**Table 1.**
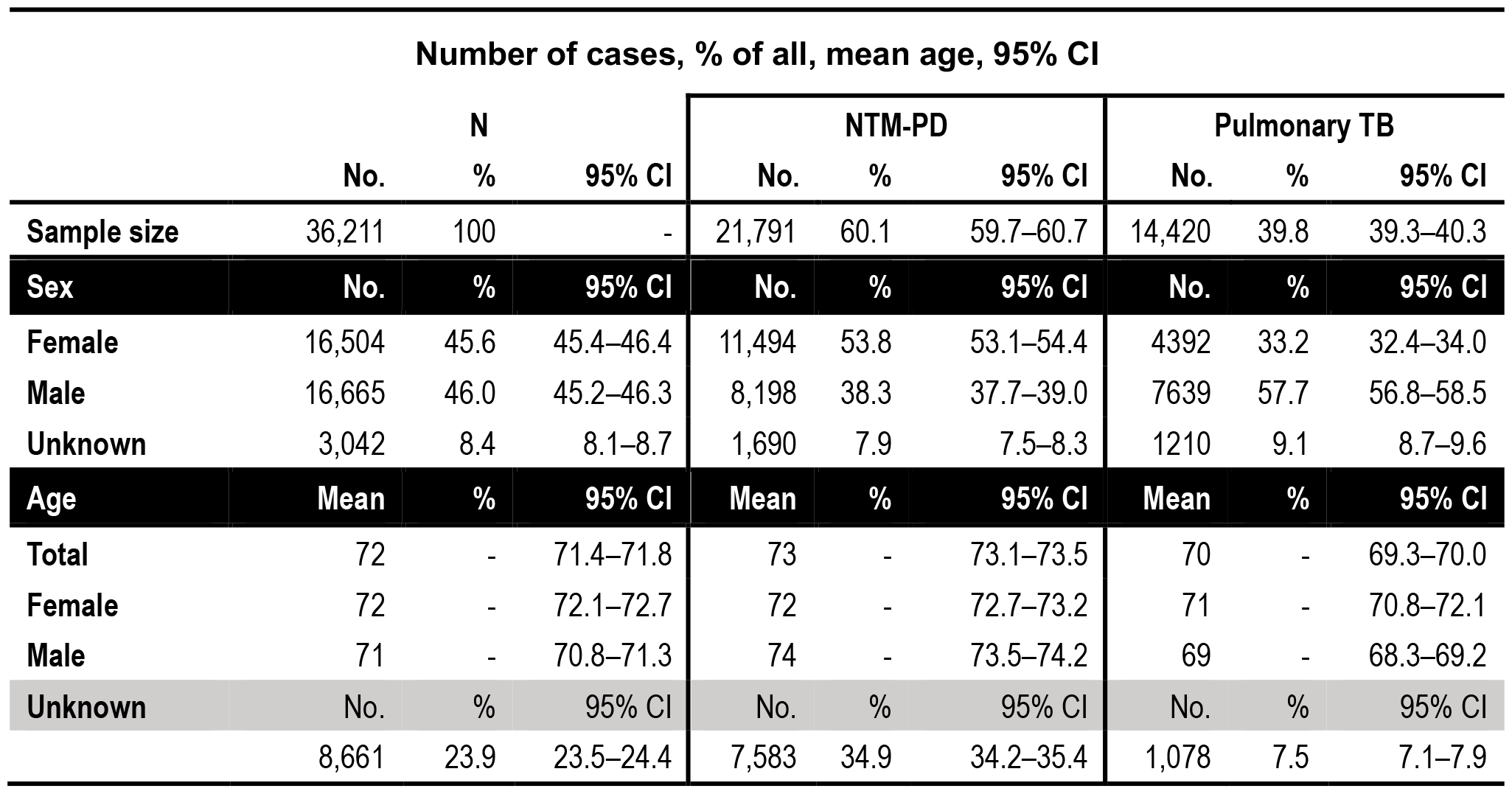
Summary of descriptive biological and bacteriological data analysed in this study.

| Number of cases, % of all, mean age, 95% CI |  |  |  |  |  |  |  |  |  |
| --- | --- | --- | --- | --- | --- | --- | --- | --- | --- |
|  | N |  |  | NTM-PD |  |  | Pulmonary TB |  |  |
|  | No. | % | 95% CI | No. | % | 95% CI | No. | % | 95% CI |
| Sample size | 36,211 | 100 | - | 21,791 | 60.1 | 59.7–60.7 | 14,420 | 39.8 | 39.3–40.3 |
| Sex | No. | % | 95% CI | No. | % | 95% CI | No. | % | 95% CI |
| Female | 16,504 | 45.6 | 45.4–46.4 | 11,494 | 53.8 | 53.1–54.4 | 4392 | 33.2 | 32.4–34.0 |
| Male | 16,665 | 46.0 | 45.2–46.3 | 8,198 | 38.3 | 37.7–39.0 | 7639 | 57.7 | 56.8–58.5 |
| Unknown | 3,042 | 8.4 | 8.1–8.7 | 1,690 | 7.9 | 7.5–8.3 | 1210 | 9.1 | 8.7–9.6 |
| Age | Mean | % | 95% CI | Mean | % | 95% CI | Mean | % | 95% CI |
| Total | 72 | - | 71.4–71.8 | 73 | - | 73.1–73.5 | 70 | - | 69.3–70.0 |
| Female | 72 | - | 72.1–72.7 | 72 | - | 72.7–73.2 | 71 | - | 70.8–72.1 |
| Male | 71 | - | 70.8–71.3 | 74 | - | 73.5–74.2 | 69 | - | 68.3–69.2 |
| Unknown | No. | % | 95% CI | No. | % | 95% CI | No. | % | 95% CI |
|  | 8,661 | 23.9 | 23.5–24.4 | 7,583 | 34.9 | 34.2–35.4 | 1,078 | 7.5 | 7.1–7.9 |

### 3.2. Estimated annual incidence of NTM-PD

The notification (incidence) rate (per 100,000) of pulmonary TB steadily decreased from 12.5 (95% CI; 12.3–12.7) in 2013 to 10.3 (95% CI; 10.1–10.5) in 2017 (P value < 0.001) (Supplementary Figure S1). In contrast, the estimated annual incidence rate of NTM-PD increased by 17.7%, from 15.8 (95% CI: 14.9–16.8) in 2013 to 19.2 (95% CI; 18.2–20.4) in 2017, with significant time-dependent trends (P value < 0.001) (Figure 2).

**Figure 2.**
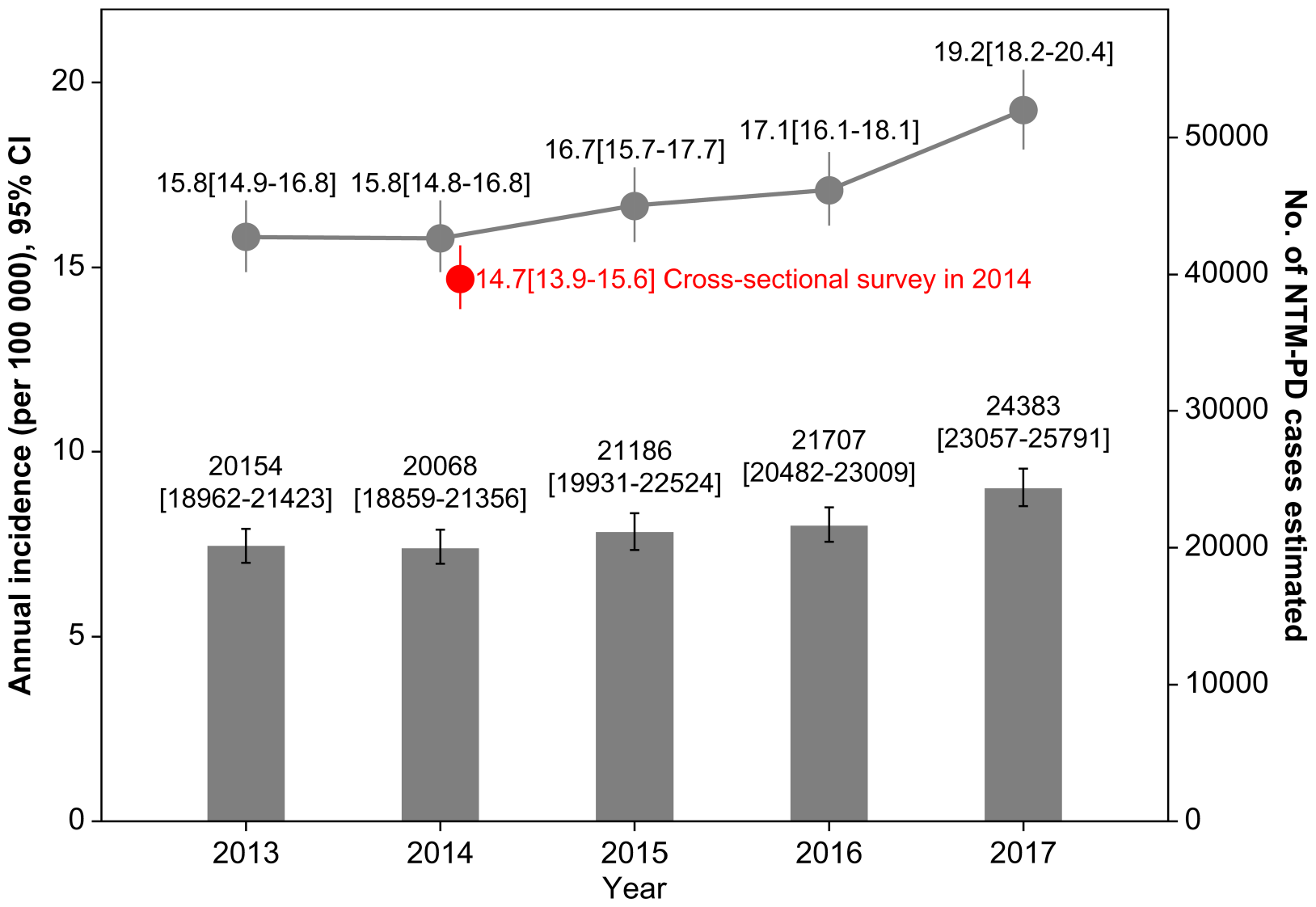
Estimated annual pulmonary diseases with non-tuberculous mycobacteria (NTM-PD) incidence and notification rates of pulmonary tuberculosis (TB) for 2013–2017. Black asterisk points show the estimated annual incidence (per 100,000) of NTM-PD, and greyish bars show the estimated number of NTM-PD cases. Solid error bars represent 95% confidence intervals assuming a binomial distribution. The notification rates of pulmonary TB were calculated by dividing the number of pulmonary TB cases by the Japanese population. The TB cases were routinely notified by health facilities and registered in the National Epidemiological Surveillance of Infectious Disease, and are shown as circles. The incidence of NTM-PD estimated by another research [5] group in 2014 is shown in red (we originally calculated the 95% confidence intervals using their [5] reported ratio, [2,652 NTM-PD cases]/[2,327 TB cases]). The mathematical formula for estimating the NTM-PD incidence is available in Supplementary Figure S2.

### 3.3. NTM species isolated from culture-positive specimens

Once *Mycobacteria* were isolated from cultures using Ogawa or MGIT methods, the species were identified using NAT, DDH, or MALDI-TOF MS analyses. Notably, MALDI-TOF MS was introduced in 2017; therefore, NAT- or DDH-tested results were mostly used for analysis. Of the 21,791 NTM-PD cases, species could be identified for 647 (39.7%), including 514 (2.04%) cases of multi-infection (Figure 3). Of the successfully identified NTM cases, 93.0% were *M. avium-intracellulare* complexes associated with *M. avium* (62.0%) and *M. intracellulare* (31.0%), followed by *M. abscessus* (2.2%) and *M. kansasii* (2.1%).

**Figure 3.**
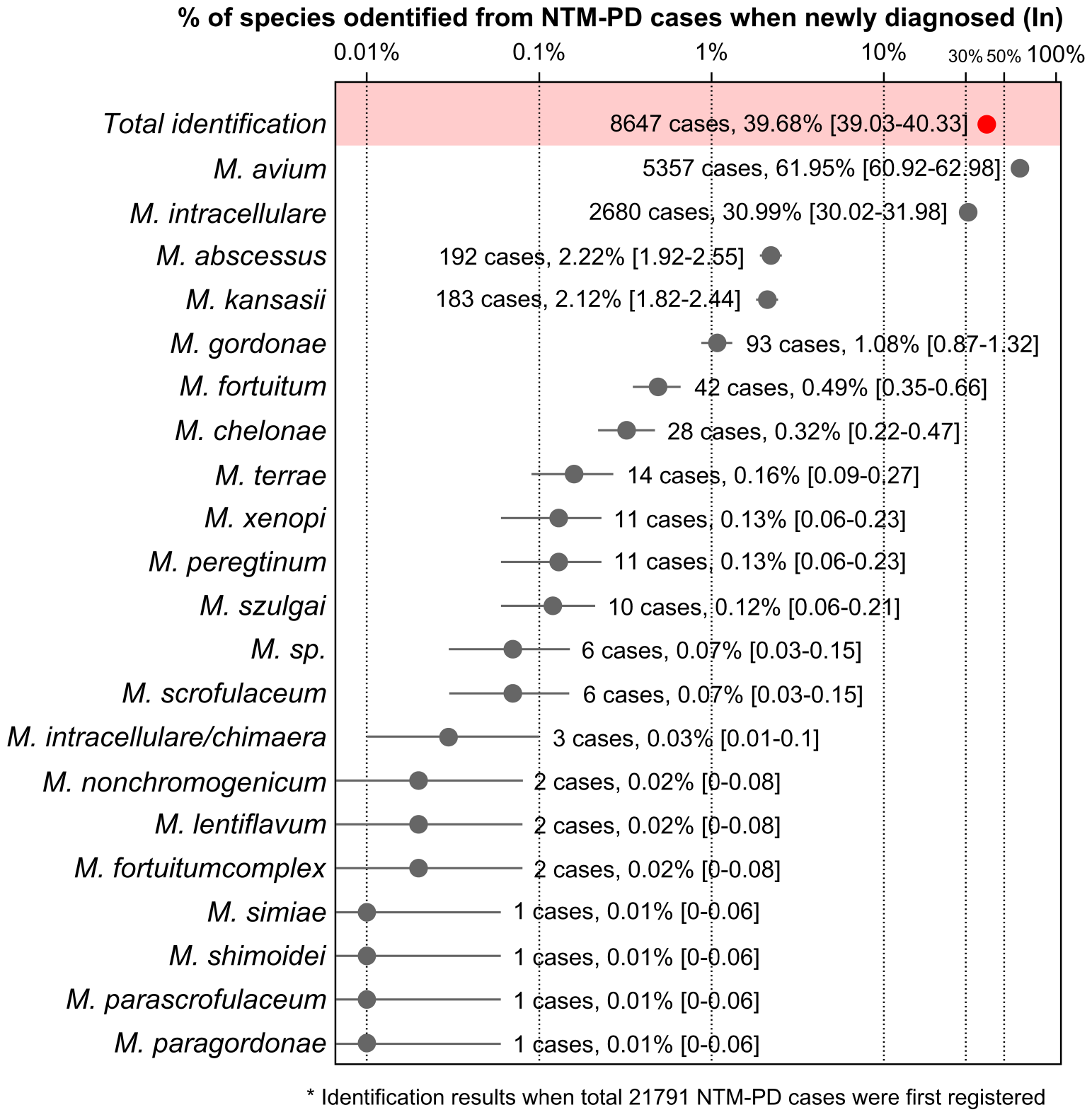
Number and percentage of *Mycobacterium* species among the 8,647 pulmonary diseases with non-tuberculous mycobacteria (NTM-PD) cases newly diagnosed with an NTM identification. Since several examinations were performed per case, multiple species were commonly isolated (note that the ‘date of first examination’ was defined as the ‘date of new diagnosis’, assuming that each NTM-PD case was caused by the species most often first detected in sequential examinations). Therefore, we described in detail the percentage and number of NTM-PD cases with successful NTM identification (95% confidence intervals based on a Poisson distribution).

### 3.4. Age dependency and gender difference in NTM-PD incidence

Of all of the NTM-PD cases (21,791) identified from 2013 to 2017, sex and age could be identified in 20,090 (8.8% missing) and 7583 cases (34.9% missing) cases, respectively. A complete dataset consisting of 13,817 patients (66.4% of all NTM-PDs) with information on both sex and age was included in the final analysis (Figure 4).

**Figure 4.**
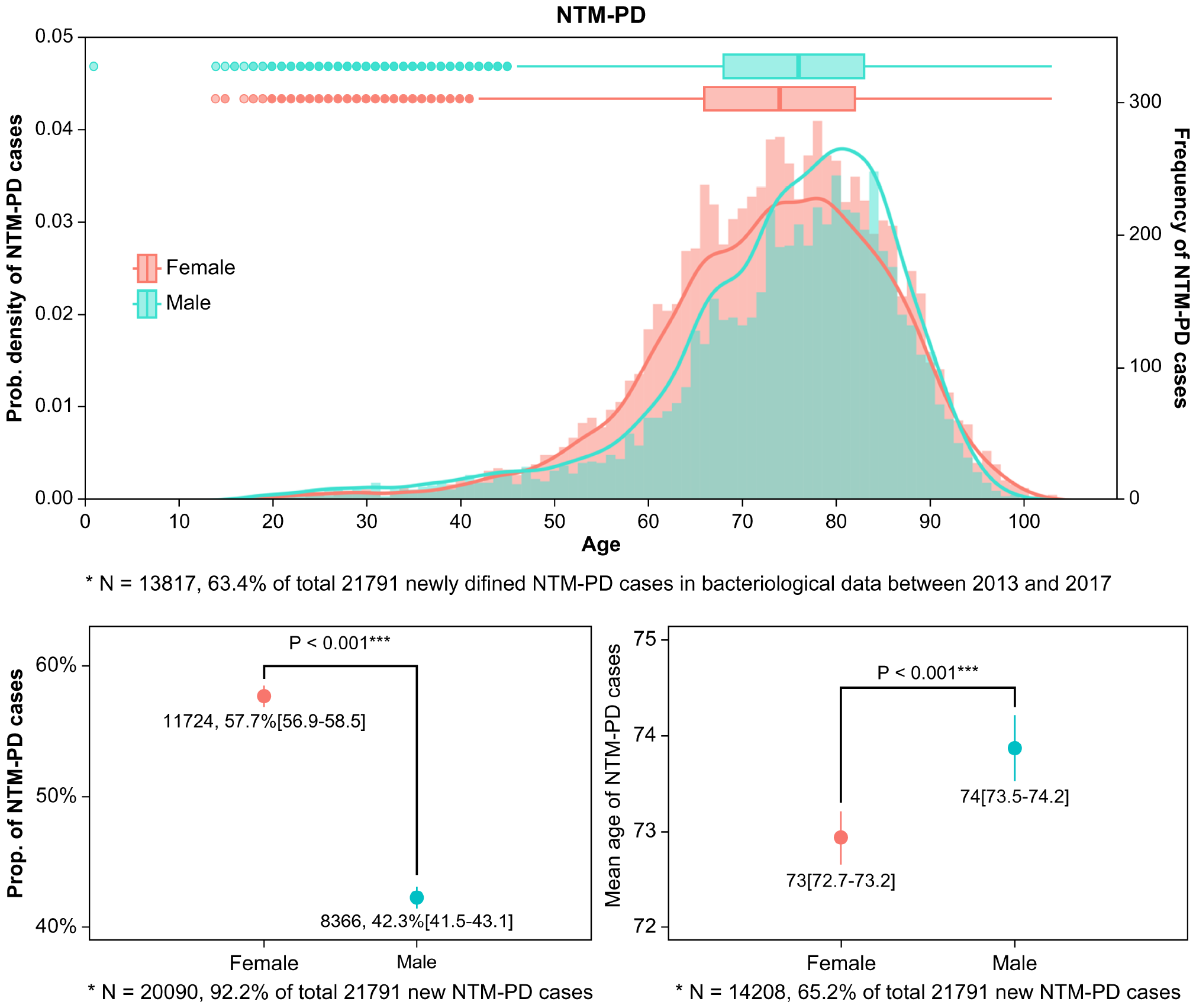
Distributions of age- and sex-specific pulmonary diseases with non-tuberculous mycobacteria (NTM-PD) cases. Summary of NTM-PD cases between 2013 and 2017 according to sex and age. In the upper panels, the two coloured lines and bars show the probability density and frequency of NTM-PDs according to sex, respectively. The proportions and mean ages of female and male patients were compared using a chi-square test. Error bars indicate 95% confidence intervals assuming Poisson (or binomial) distribution.

Females accounted for 57.7% (56.9–58.5) of all NTM-PD cases, and their proportion was significantly greater than that of males (42.3% (41.5–43.1), P < 0.001, Figure 4; left bottom). The mean age of patients with NTM-PDs was significantly less in females than in males, 73 (72.7–73.2) and 74 (73.5–74.2), respectively (P < 0.001, Figure 4; right bottom). NTM-PDs occurred most frequently in males in their 70s and in females in their 80s (Figure 4, upper panel). Moreover, between the ages of late 40s and early 70s, NTM-PDs occurred more predominantly in females, after which cases in males dominated. A significant age-dependent trend was observed (P < 0.001).

## 4. Discussion

We integrated bacteriological data from two private laboratories to bacteriologically define NTM-PD and pulmonary TB cases. We then sequentially estimated the annual NTM-PD incidence rates per 100,000 individuals from 2013 to 2017, to clarify the dynamics of NTM-PD epidemics. We used a survey method for estimating NTM-PD incidence, which was previously implemented in independent studies [8, 12]. Descriptive analyses of pulmonary TB cases revealed a similarity in the distribution between the bacteriological data and national TB surveillance data, thereby supporting the validity of this methodology. In our study, the annual incidence of NTM-PDs exhibited an upward trend since 2013, peaking at 19.2 (per 100,000) in 2017. The age- and sex-specific distribution of NTM-PD cases highlighted a predominance of middle-aged females, with a shift to male predominance in individuals aged ≥80 years. *Mycobacterium abscessus* emerged as the second most common species, surpassing *M. kansasii* for the first time in Japan.

A cross-sectional survey in 2014 [5] estimated the incidence of NTM-PD (per 100,000) at 14.7, a 2.6-fold increase from the 5.7 estimated in a previous survey [13] in 2007. Similarly, we found a continuous upward trend, with a 17.7% increase in the NTM-PD incidence between 2013 and 2017, resulting in the NTM-PD incidence of 15.8 (per 100,000) in 2014, which was mostly consistent with the results of the cross-sectional survey. However, these results were not directly comparable since a uniform formulation was not used. While we estimated the NTM-PD incidence, using the same method (NTM-PD / pulmonary TB; Supplementary Figure S2) as the survey [13] in 2007 or earlier, the survey in 2014 [5] also used the same method but with a slightly modified formulation (NTM-PD / all forms of TB). The use of a non-uniform formulation could affect external consistency. Moreover, the monthly TB notification rates previously used should have massive seasonal variations, which might result in over- or underestimation of NTM-PD incidence. Additionally, the use of a shorter study period spanning only two months was not sufficient to handle bias correction in the study results. Further well-designed studies are warranted to confirm the validity of these findings. Pulmonary TB and NTM-PDs are often diagnosed using a similar approach based on bacteriological examinations using respiratory specimens; thus, we propose the use of annual pulmonary TB notification rates to estimate NTM-PD incidence in addition to a uniform formula (Supplementary Figure S2).

Analysis of the five-year incidence of NTM-PDs (per 100,000) revealed that female cases have increasingly become more predominant than male cases, with this trend being particularly evident among females in their 40s, 70s, and especially in their 60s. Conversely, male cases were more prevalent in the 80s and older. Similar epidemiological trends in NTM-PD have been observed in studies analysing national health insurance claims for nearly the entire Japanese population, supporting the representativeness of this sample population, albeit on a laboratory basis [2]. Notably, another study [14] reported that the proportion of underlying lung diseases, such as previous TB and chronic obstructive pulmonary disease, increased with age in male patients, whereas no distinct trend was observed in female patients. A sex disparity in the NTM-PD incidence was evident within the population at risk of sex-specific underlying conditions.

To our knowledge, this is the first study to demonstrate that the prevalence of *M. abscessus* among NTM-PD cases surpassed that of *M. kansasii*. Notably, *M. abscessus* is the second most common species in East Asia, including Korea and Taiwan [15-19]. Furthermore, the Kyushu and Okinawa regions, located in southwestern Japan and in proximity to these East Asian countries (South Korea and Taiwan), exhibit a higher prevalence of the NTM infection with *M. abscessus* species. A rising trend in the NTM-PD with *M. abscessus* in the east Japan has been documented as well [14]. Therefore, emphasising epidemiological research to reveal a species-specific geographical heterogeneity in the East Asia is imperative. Moreover, as NTM is an environmental bacterium that exists anywhere, it is important to identify impact factors at the global (e.g. climate change, geo-space) to household (e.g. water system and equipment, soils and minerals surrounding the living space) level, and to propose public health approaches against NTM-disease burden [3] from the aspect of ‘One Health’.

This study has several limitations. First, there could be sampling bias, resulting in an overestimation of NTM incidence. We used information from bacteriological examinations intended to diagnose infectious diseases with mycobacteria; thus, the chance of isolating NTM and defining NTM-PDs in our study population could be greater than that in random sampling from a general population. Since both NTM-PD and pulmonary TB cases were more likely to be observed in our study population, we strictly defined the respiratory diseases associated with *M. tuberculosis* isolated from specimens (respiratory or gastric fluid) specified for TB diagnosis as pulmonary TB. Furthermore, among the pulmonary TB cases, the age and sex distributions of our study population mostly reflected those of pulmonary TB notifications in Japan, as previously described. Consequently, we were less likely to observe a significant difference between the ratio of NTM-PD cases to pulmonary TB cases within the general population and that within our observation for use in the uniform formulation (Supplementary Figure S2) to estimate the NTM incidence rate, ensuring external validity in our study. Further, since there could be potential NTM-PD cases that were not diagnosed and subsequently recovered, similar to that of TB cases, the NTM-PD incidence could indeed be underestimated rather than overestimated, as suggested previously [20].

As indicated in this study, the centralised integration of bacteriological data in private microbiology laboratories shows enormous potential in the establishment of a nationwide NTM surveillance system, where individual clinical status is connected to each piece of bacteriological information. The laboratory-based surveillance system not only enables continuous monitoring of epidemic dynamics of NTM without time-consuming surveys, but it also accelerates the development of the NTM epidemiology research field.

## 5. Conclusion

In this study, we developed a preliminary laboratory-based surveillance system for respiratory infections with NTM to integrate bacteriological information from collaborative private microbiology laboratories along with a computer algorithm for bacteriologically case-defined criteria. We successfully implemented sequential estimates of NTM-PD incidence between 2013 and 2017, and statistically revealed time- and age-dependent trends and sex differences in the incidence of NTM-PDs. These findings, despite being laboratory-based, are extrapolatable to the general population in Japan and support our system as a viable alternative to the nationwide surveillance system for NTM. This study is the first step in establishing an NTM surveillance system in Japan. Further studies are needed to involve more private laboratories to cover a larger population, and a nationwide cross-sectional survey is required to validate the effectiveness of our surveillance system, despite potential intermittent use. Our current findings and future studies will contribute to NTM epidemiology, further elucidating areas that are still unexplored and mitigating the challenges associated with NTM-PDs.

## Data Availability

All data generated in this study are essentially unavailable.

## Declaration of competing interest

The authors declare that this study was conducted in the absence of any commercial or financial relationships that could be construed as potential conflicts of interest.

## Funding

This research was supported by the Research Program on Emerging and Reemerging Infectious Diseases of the Japan Agency for Medical Research and Development (AMED; 23fk0108673h0801).

## CRediT authorship contribution statement

**Yuko Hamaguchi**:Conceptualisation, Methodology, Software, Formal analysis, Validation, Data curation, Writing the original draft, Writing the review and editing, and Visualisation. **Kozo Morimoto**: Conceptualisation, Methodology, Formal analysis, Resources, Writing – review and editing, Supervision, Project administration, and funding acquisition. **Satoshi Mitarai**: Conceptualisation, Methodology, Formal analysis, Resources, Writing, review and editing, Supervision, Project administration.

## Acknowledgements

The microbiological data used in this study were coordinated by SRL, Inc. (Kazushi Uematsu and Jun Sokunaga), LSI Medience Corporation (Akiko Nakata and Shunsuke Shibuya), and BML, Inc. (Takayuki Tomii).

## Notes

### Competing Interest Statement

The authors have declared no competing interest.

### Author Declarations

This study was approved by the Ethics Committee of Fukujuji Hospital (approval #19046)

### Summary of Updates

There has been no revision at the moment.

