## Supplementary Materials for "Laboratory-based surveillance of non-tuberculous mycobacterial pulmonary disease in Japan"

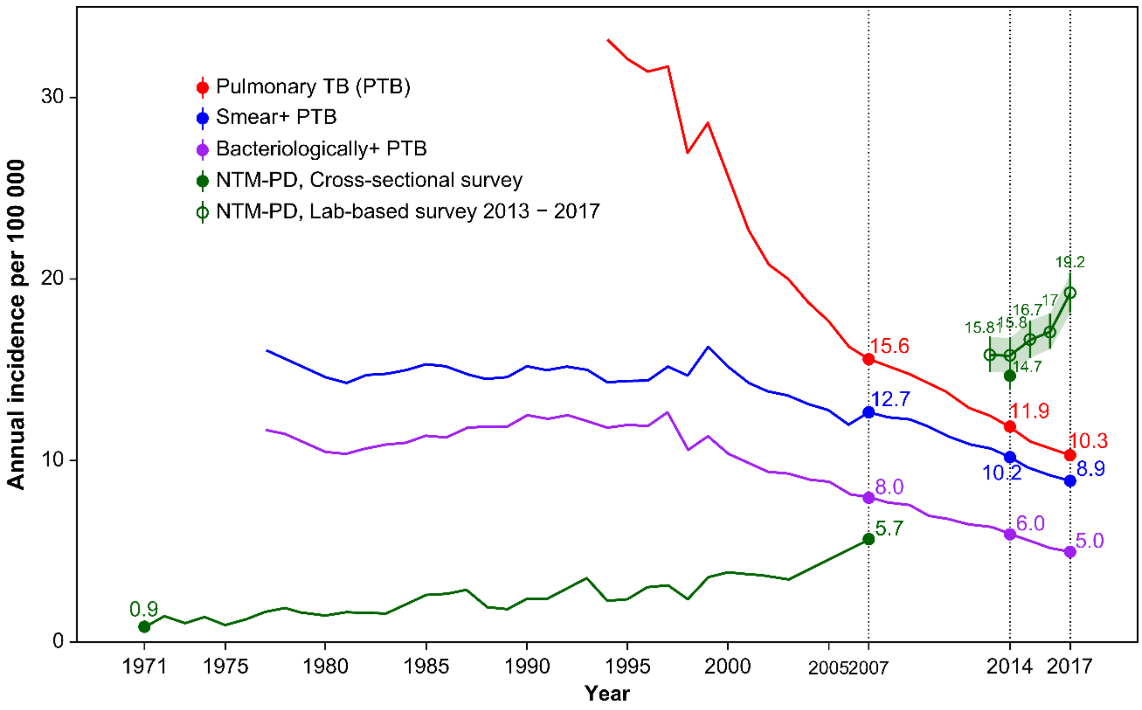


**Supplementary Figure S1**. Incidence rates of pulmonary tuberculosis (TB) and pulmonary diseases with non-tuberculous mycobacteria (NTM-PD) since 1971

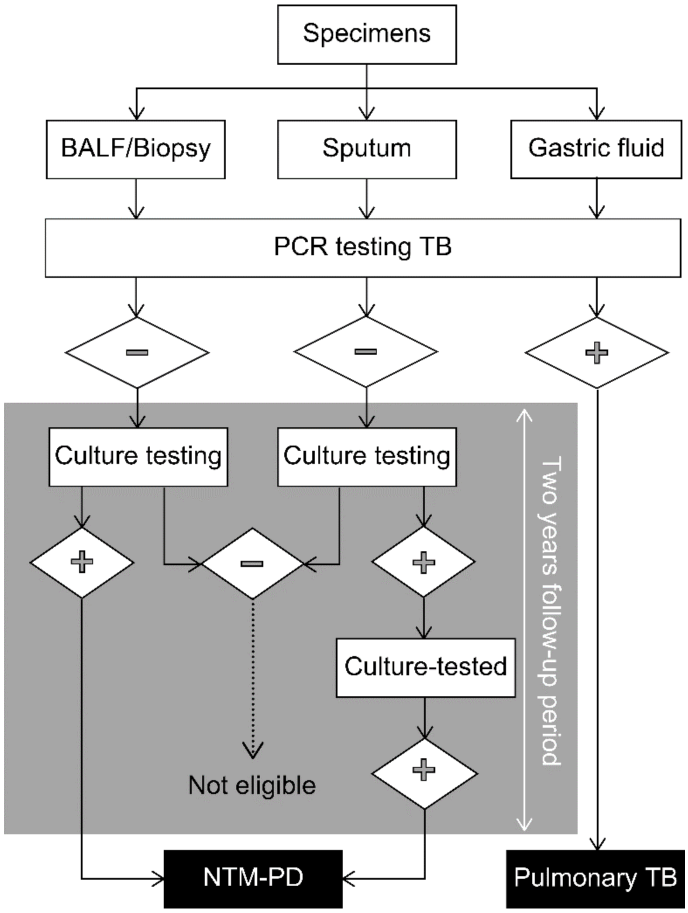


**Supplementary Figure S2**. Algorithm of bacteriological criterion defining pulmonary diseases with non-tuberculous mycobacteria (NTM-PD) and pulmonary tuberculosis (TB)

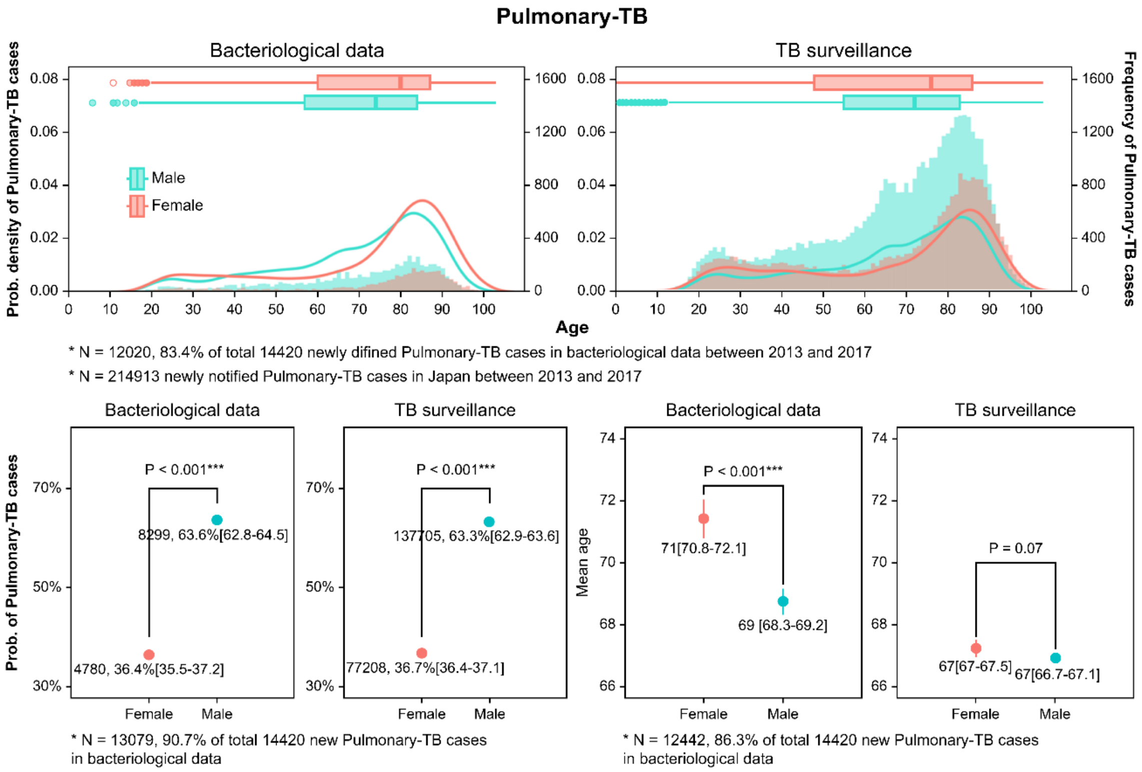


**Supplementary Figure S3**. Distribution of pulmonary tuberculosis (PTB) patients by sex and age and comparison of tuberculosis surveillance data with bacteriological data. PTB reported in the surveillance system and PTB defined in bacteriological data between 2013–2017 are shown according to each sex and age group. In the upper panels, the two coloured lines and bars show the probability density and frequency of pulmonary diseases with non-tuberculous mycobacteria (NTM-PDs) according to sex, respectively. The proportion and mean age of female and male patients were compared using the chi-square test and Wilcoxon rank-sum test. Error bars indicate 95% confidence intervals assuming a Poisson (or binomial) distribution.

**Supplementary Table S1**. Formulation of a unified indicator to estimate pulmonary diseases with non-tuberculous mycobacteria (NTM-PD) incidence

| | |
|---|---|
| $TB_{case}$ | Number of pulmonary tuberculosis (TB) cases in a population (national surveillance-based) |
| $\widetilde{TB_{case}}$ | Number of pulmonary TB cases in a sample (cases in which respiratory specimens tested polymerase chain reaction (PCR)-positive in private laboratories for our study) |
| $TB_{coverage}$ | Pulmonary-TB case coverage, defined as the proportion of pulmonary TB cases in a sample to those in a population<br>$TB_{coverage} = \widetilde{TB_{case}} / TB_{case} \cdots (1)$ |
| $NTM_{case}$ | Number of NTM cases in a population |
| $\widetilde{NTM_{case}}$ | Number of NTM-PD cases in a sample (cases in which conditions satisfy bacteriological NTM-PD case definition in our study) |
| $NTM_{coverage}$ | NTM-PD case coverage, defined as the proportion of NTM-PD cases in a sample to those in a population<br>$NTM_{coverage} = \widetilde{NTM_{case}} / NTM_{case} \cdots (2)$ |
| $TB_{rate}$ | TB incidence rate, defined as the proportion of TB cases in a population within a year (commonly using a unit of $10^n$)<br>$TB_{rate} = \left[ TB_{case} / Number\ of\ population \right] \times 100{,}000 \cdots (3)$ |
| $NTM_{rate}$ | NTM-PD incidence rate, defined as the proportion of NTM-PD cases in a sample within a year<br>$NTM_{rate} = \left[ NTM_{case} / Number\ of\ population \right] \times 100{,}000 \cdots (4)$ |

By approximating the pulmonary TB coverage to the NTM-PD coverage in private laboratories (a sample), we derived a formula from (2).

$NTM_{coverage} \approx TB_{coverage} = \widetilde{NTM_{case}} / NTM_{case}$.

Then,

$NTM_{case} = \widetilde{NTM_{case}} / TB_{coverage} \cdots (6)$.

(6) was modified using (1) as follows:

$$NTM_{case} = \widetilde{NTM_{case}} \Big/ \left[ \widetilde{TB_{case}} / TB_{case} \right].$$

$$NTM_{case} = \left[ \widetilde{NTM_{case}} / \widetilde{TB_{case}} \right] \times TB_{case} \cdots (7).$$

Substituting (7) into (4) yielded

$$NTM_{rate} = \left[ \left[ \widetilde{NTM_{case}} / \widetilde{TB_{case}} \right] \times TB_{case} \Big/ Number\ of\ population \right] \times 100{,}000.$$

The above formulation was then simplified by modifying it using (3) as follows:

$$NTM_{rate} = \left[ \widetilde{NTM_{case}} / \widetilde{TB_{case}} \right] \times TB_{rate} \cdots (8).$$

(8) indicates the annual NTM-PD incidence rate obtained using a simple formula in which the ratio of NTM-PD cases to pulmonary TB cases in the sample (cases in private laboratories) was multiplied by the annual pulmonary TB incidence rate in the population.
